# Acceptability of a community health worker- led health literacy intervention on lifestyle modification among hypertensive and diabetes patients in the City of Harare, Zimbabwe

**DOI:** 10.1101/2024.07.10.24310219

**Authors:** Nyaradzai Arster Katena, Shepherd Shamu, Golden Tafadzwa Fana, Evans Dewa, Admire Dombojena, Simbarashe Rusakaniko

**Author notes:** **Corresponding Author** Nyaradzai A. Katena University of Zimbabwe Faculty of Medicine & Health Sciences, Department of Global Public Health &Family Medicine P.O Box A178, Avondale Harare Zimbabwe.

## Abstract

Deploying community health workers (CHWs) is a vital strategy to improve health at a community level in low- and middle-income countries. Whilst there is substantial evidence on the effectiveness of CHWs interventions, there is a need for more research on the mechanisms through which these interventions work. Understanding the acceptability of these interventions is one way of assessing the mechanisms through which they work. This article examines the acceptability of a community health worker-led health literacy intervention on lifestyle modification among hypertensive and diabetes patients based on the perspectives of the CHWs, community nurses and diabetes and hypertensive patients. A qualitative study was imbedded within a cluster randomized trial to assess the effectiveness community health worker-led health literacy intervention on lifestyle modification among hypertensive and diabetes patients in the City of Harare, Zimbabwe. Data were gathered through semi-structured interviews with 3 community health nurses and 15 diabetes and hypertension patients as well as 2 focus group discussions with CHWs. Data were analyzed manually using the thematic analysis method. There was consensus that the intervention had many benefits amongst CHWs and community nurses. However, among patients, there were mixed perceptions regarding the benefits of the intervention. The main challenges that were mentioned by CHWs include resistance to advice by patients, insufficient resources, and lack of acceptance at some of the patient’s homes. All participants believed the intervention was acceptable. Our study provides vital information that should be considered in upscaling CHW led interventions.

## INTRODUCTION

Deploying community health workers (CHWs) is a vital strategy to improve health at a community level in low- and middle-income countries (LMICs) [1–4]. Community health workers are voluntary public health workers who play a crucial role of primary and tertiary prevention of diseases[5]. Implementation of public health interventions through CHWs can facilitate the linkage between communities and health services and thus ensure community participation. For many years, CHWs in LMICs have been providing community-based care, but their efforts have been focused on home-based caring for people living with the HIV and TB, with little being done around non-communicable diseases[5]. This is despite an increase in the burden of NCDs in LMICs over the recent years. Worldwide, NCDs kill 41 million people each year, which is equivalent to 74% of all deaths globally[6,7]. Each year, 17 million people die from a NCD before the age of 70 and 86% of these premature deaths occur in LMICs . Overall, 77% of all NCDs deaths are in LMICs. Amongst all NCDs, cardiovascular diseases account for most NCD deaths, with 17.9 million deaths annually[6,7].

The contribution of CHWs to infectious chronic diseases care in many LMICs suggests they could play a significant role in NCDs management in these countries. Management of NCDs in these countries is often poor with health systems struggling with several challenges that include staff shortages[8]. Hence involvement of CHWs in the management of NCDs is important. In the countries where the CHWs participated in the management of NCDs, their roles had included education and awareness, screening and monitoring, support for medication and lifestyle modification adherence, referral to healthcare professionals and advocating for healthier environments[5,9,10].

Diabetes and hypertension are amongst the most prevalent NCDs worldwide and CHW-led interventions for these conditions are increasing becoming popular in LMICs[11]. Overall, CHWs provide additional workforce for the management and care of hypertension and diabetes patients where resources are limited and allow for task shifting to alleviate overstretched health systems [12,13]. Several studies have reported positive outcomes for CHWs interventions such as reduced cardiovascular risk, improved medication adherence, improved lifestyle modification and improved quality of life[3,14–18]. Community health workers can play a beneficial role in improving health outcomes among diabetes and hypertensive patients, while improving equity in health care delivery[19] (Jeet et al. 2017)]. As a result, it is increasingly recommended that services be offered outside of health facilities in communities, to address the rising burden of hypertension and diabetes.

There are several studies that indicate that the global increase in the burden of hypertension and diabetes has seen a growing use of CHWs to support patients on treatment and this has resulted in good clinical outcomes. In a scoping review of 54 articles on the role of community health workers in type 2 diabetes mellitus self-management, it was concluded that CHWs play several roles, including structured education, ongoing support and health system advocacy [20]. Another review on studies evaluating the effectiveness of community-based interventions for prevention of Type 2 diabetes in LMICs also confirmed the successful use of CHWs in management of type 2 diabetes [21]. Gyawali and Kallestrup recommended that more evidence from randomized controlled trials on culturally tailored, community-based interventions is needed to compare findings and test implementation in practice[21]. In a participatory qualitative research project carried out by Chimberengwa and Naidoo, it was concluded that in management of hypertension, CHWs were a key link between the community and the formal health delivery care system [18].

Whilst there is substantial evidence that CHW programs improve a range of health outcomes, these benefits tend to reduce or disappear when CHW programs are scaled up. One of the reasons for the reduction is low uptake of the interventions. Low uptake of public health interventions can be a result of poor acceptability[22]. Acceptability has increasingly become a key consideration in the design and implementation of public health interventions[22] . Successful implementation and uptake of a public health intervention depend on acceptability of the intervention by health care providers, communities, and the patients. For instance, if an intervention is considered acceptable by patients, they are more likely to adhere to treatment recommendations and thus benefit from improved clinical outcome. Likewise, if healthcare providers consider an intervention as acceptable, they are more likely to deliver it as intended, which also enhances the effectiveness of the intervention.

In Zimbabwe, CHWs provide community-based care, but for years, their efforts have been focused on home-based care for people with HIV and TB, with little done around NCDs[23]. This shows the need to continuously come up with effective CHW-led interventions for management common NCDs. This paper reports on a qualitative study to assess the acceptability of a community health worker-led health literacy intervention on lifestyle modification among hypertensive and diabetes patients in the City of Harare, Zimbabwe. The study was imbedded in a cluster randomised trial to assess the effectiveness of the intervention.

## METHODS

### Ethics statement

Ethics approval was granted by the Joint Research Ethics Committee for University of Zimbabwe Faculty of Medicine & Health Sciences and the Parirenyatwa Group of Hospitals (Ref: JREC 339/ 2022) as well as the Medical Research Council of Zimbabwe (Ref: MRCZ/A/3059). Formal written Informed consent was obtained from participants before data collection.

### Study setting

The study was conducted in 3 City of Harare primary health care clinics, which were in the intervention arm of the cluster randomised controlled trial to assess the effectiveness of the intervention. The 3 health facilities are part of of 39 health facilities (12 polyclinics, six family health services clinics, 15 satellite clinics and two infectious diseases hospitals) in the city. The 3 facilities offer comprehensive primary health care services, which include curative services, care of chronic patients, maternal and child health services, human immune-deficiency(HIV) prevention services and community health services. These clinics are in the high-density suburbs of the city, with each clinic catering for a population of 80 000 -100 000 people, who are mostly socio-economically disadvantaged.

### Study design

Our study employed a qualitative design The qualitative study was imbedded in an ongoing cluster randomized trial. The aim of the cluster randomized controlled trial was to assess the effectiveness of a community health worker–led health literacy intervention on lifestyle modification among patients with hypertension and diabetes, which was designed by the principal investigator (NAK).

### Conceptual Framework

Acceptability was assessed from the perspectives of the CHWs, community nurses and patients, using a conceptual framework that we adapted from 3 most used theoretical frameworks for acceptability; the Acceptability Framework by Sekhon etal, 2017; the Theoretical Framework for acceptability-TIFA by Sallis etal 2019 and the Acceptability Model by Sidani et al 2015[22]. Based on these three models, we assessed five components of acceptability were assessed, as shown in Table 1 below:

**Table 1:** Acceptability constructs used

| Acceptability construct | Definition |
| --- | --- |
| Perceived effectiveness | Beliefs about the intervention’s ability to achieve its intended goals |
| Perceived benefits | Beliefs about the advantages and positive outcomes of the intervention |
| Perceived barriers | Beliefs about the obstacles, challenges or negative consequences of the intervention |
| Perceived feasibility | Beliefs about the practicality and ease of implementing the intervention |
| Perceived compatibility and ethicality | Beliefs about how well the intervention aligns with existing values, beliefs, norms and practices. |

### Description of the community health worker -led health literacy intervention for lifestyle modification among hypertensive and diabetes patients

Participants in the intervention arm of the cluster randomised controlled trial received a simple health literacy intervention on lifestyle modification which was delivered by trained community health workers. The intervention consisted of four face-to-face interactive educational sessions and monthly support visits and the content of the educational sessions were based on the constructs of the Health Belief Model. The intervention was implemented over a period of six months. A protocol for the cluster randomised controlled trial that contains full details of the intervention the implementation has been published in the JMIR research protocols journal[24].

### Study participants

A total of 43 participants participated in the study. We interviewed 3 community health nurses, who are the supervisors of CHWs and 25 patients (6-8 patients per clinic). A total of 15 CHWs who implemented the intervention also participated in three focus group discussions (7 participants per group) .

### Data Collection

Data was collected during the implementation of the intervention (a month after the start of the implementation up to to the end of the implementation), during the period 01 August 2023-31 January 2024. The principal researcher (NAK) and 2 trained research assistants (ED and AD) were involved in the data collection and thematic data analysis. In-depth interviews (IDIs) were held with community health nurses and patients, whilst focus group discussions (FGDs) were held with the CHWs. All the FGDs and interviews were audio recorded, using the researchers’ smart phones.

We used a guide (Supporting file) that was in the vernacular language (ChiShona) to conduct the interviews and the discussions. The questions were based on our conceptual framework, described earlier in this section. Examples of questions that were included in the guides were (1) what are the main challenges are you met in implementing the intervention? (2) what are the benefits of implementing this intervention? (3) how did this intervention help you? (4) what did you like about the intervention? and (5) what did you not like about the intervention? In-depth interviews with community health nurses were conducted in a private space at the local clinic, whilst the interviews with patients were conducted at their homes. Focus group discussions with CHWs were conducted at the clinics.

### Data analysis

The thematic analysis method was used to analyze the data, in accordance with the six steps described by Braun and Clarke[25] and following a deductive-inductive approach with a pre-existing framework that was based on our conceptual framework.

The three researchers who were involved in the analysis (NAK, ED and AD) listened to all recordings and deductively coded all transcripts separately using the coding sheet derived from the analytical framework. The principal investigator (NAK) then compared the sets of coded transcripts noting differences in coding. The differences were subsequently discussed and resolved by consensus.

## RESULTS

### Characteristics of study participants

Table 2 below shows the characteristics of participants for the IDIs and FGDs

**Table 2:**
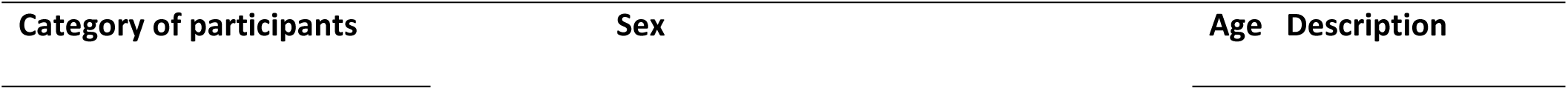
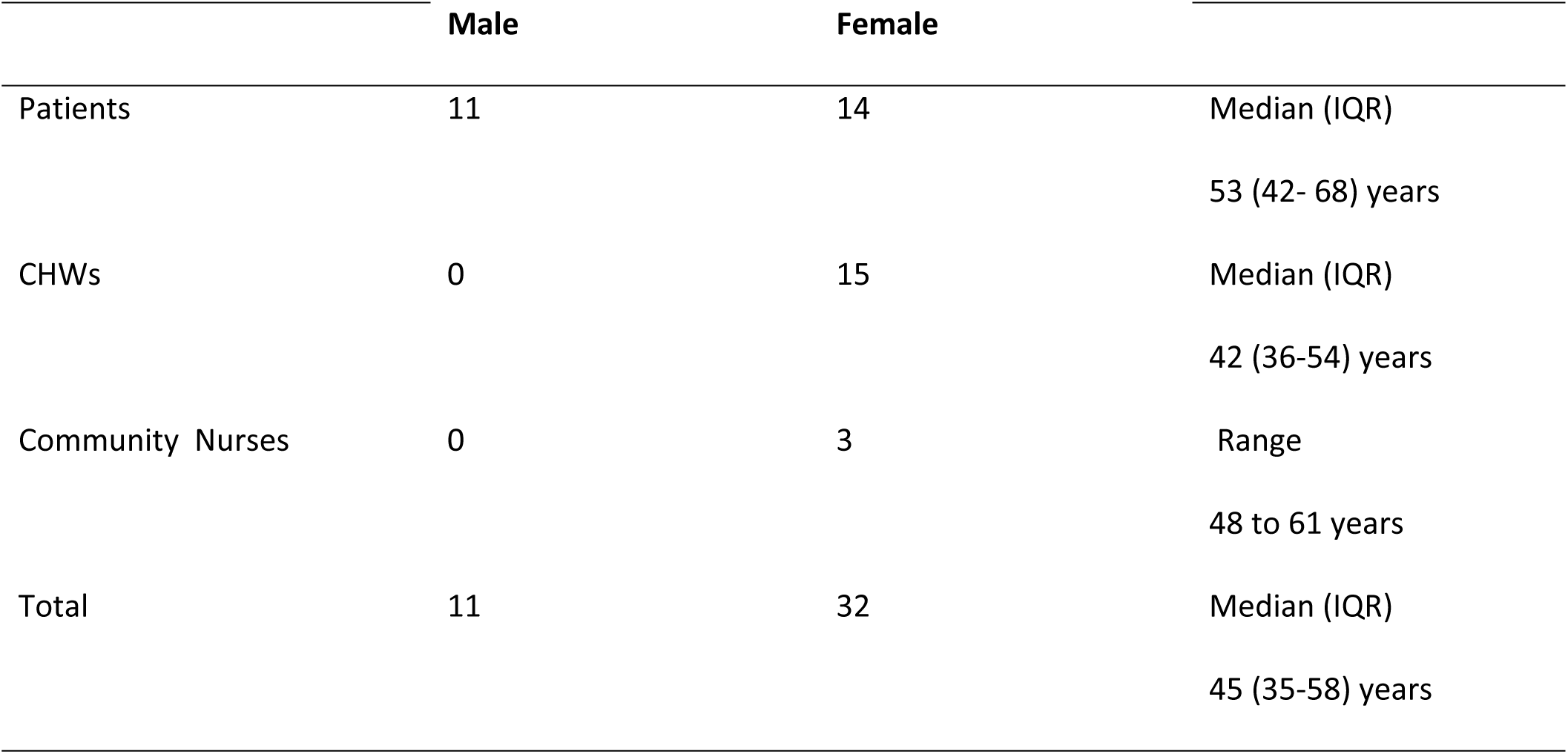
characteristics of study participants

### Participants’ Perceptions on our Intervention

Our study revealed the 5 broad themes of acceptability under the conceptual framework. There were no additional themes that were derived from the analysis . All the themes, subthemes and supporting verbatim of study are presented in Table 3.

**Table 3:**
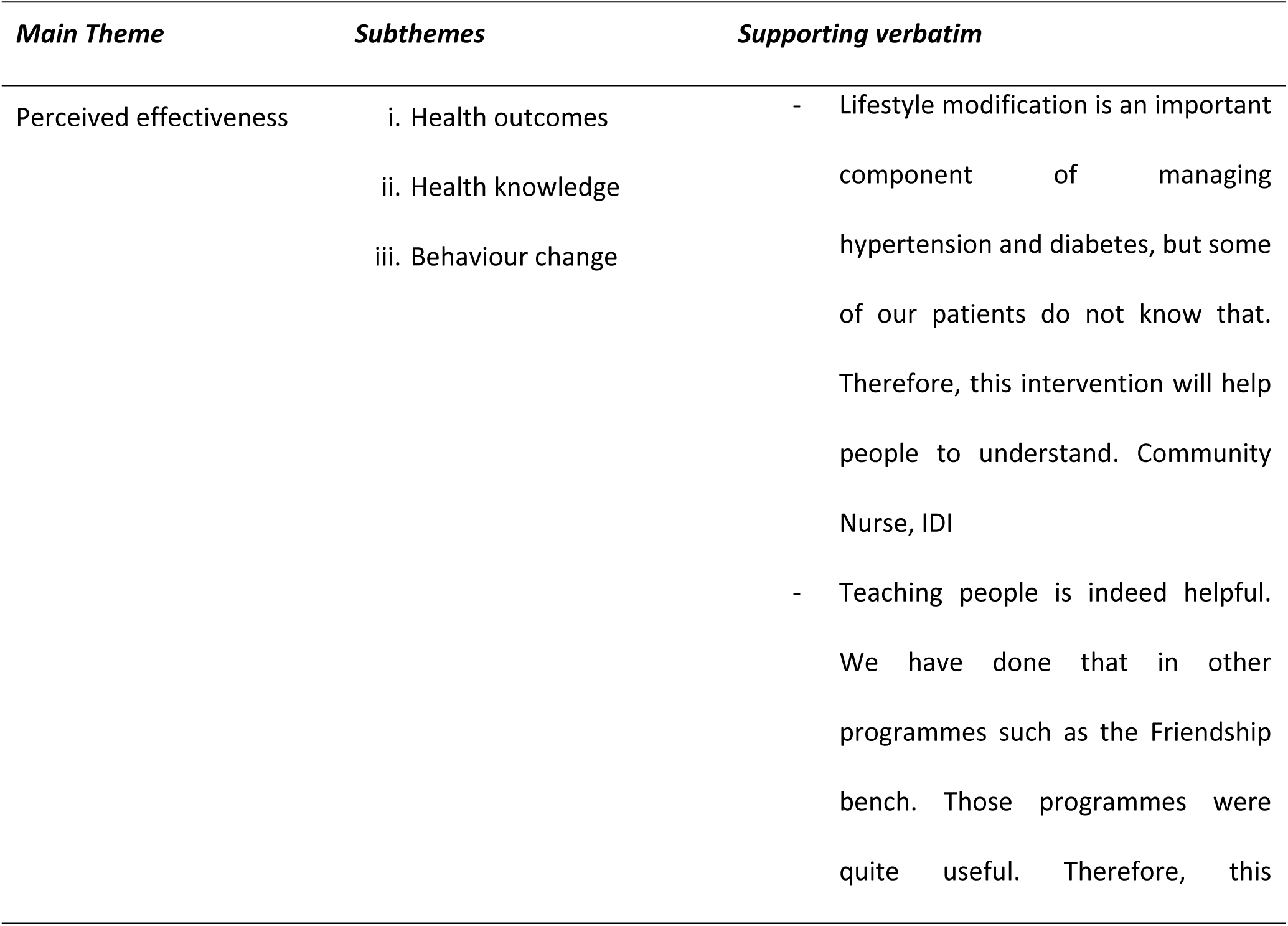
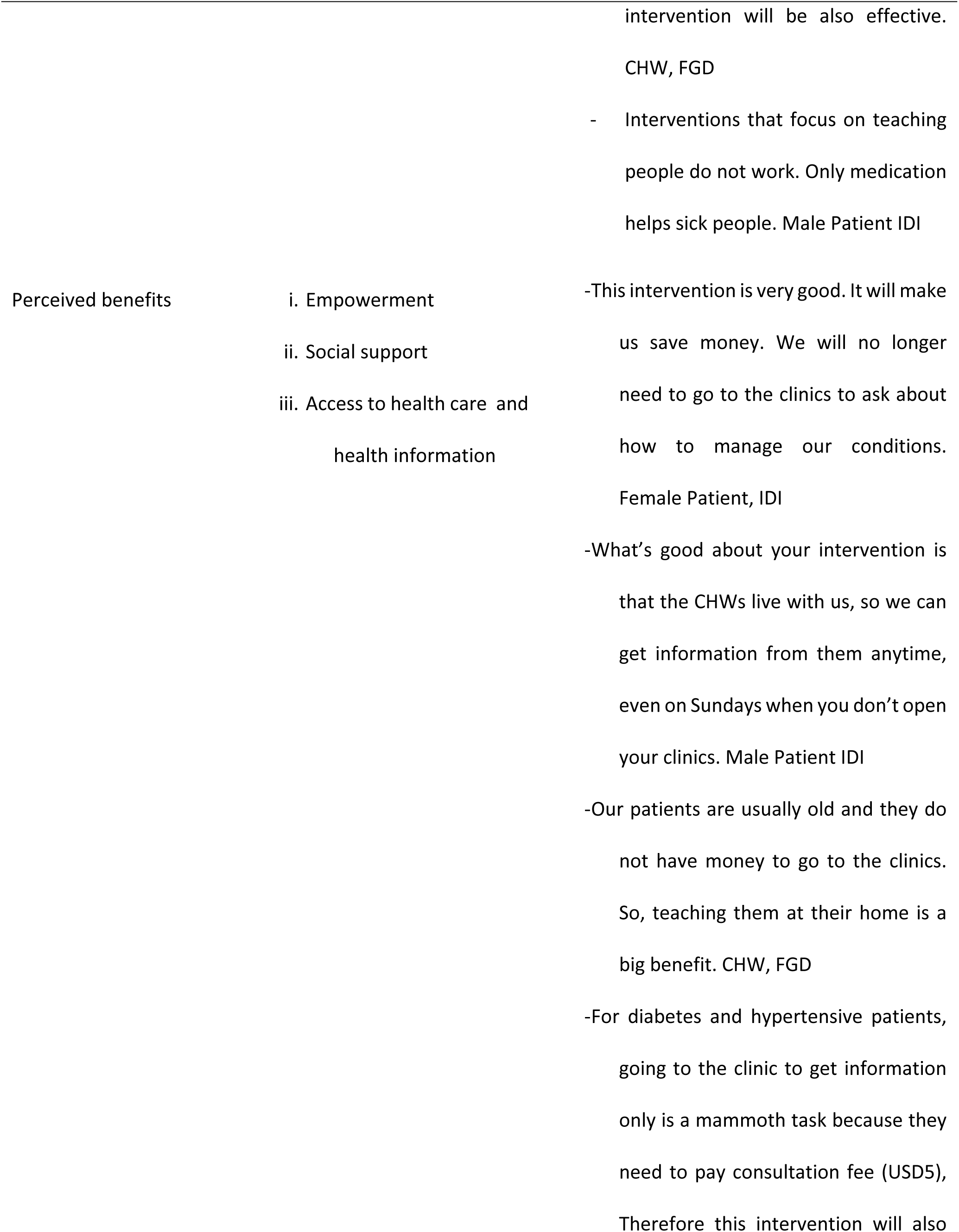
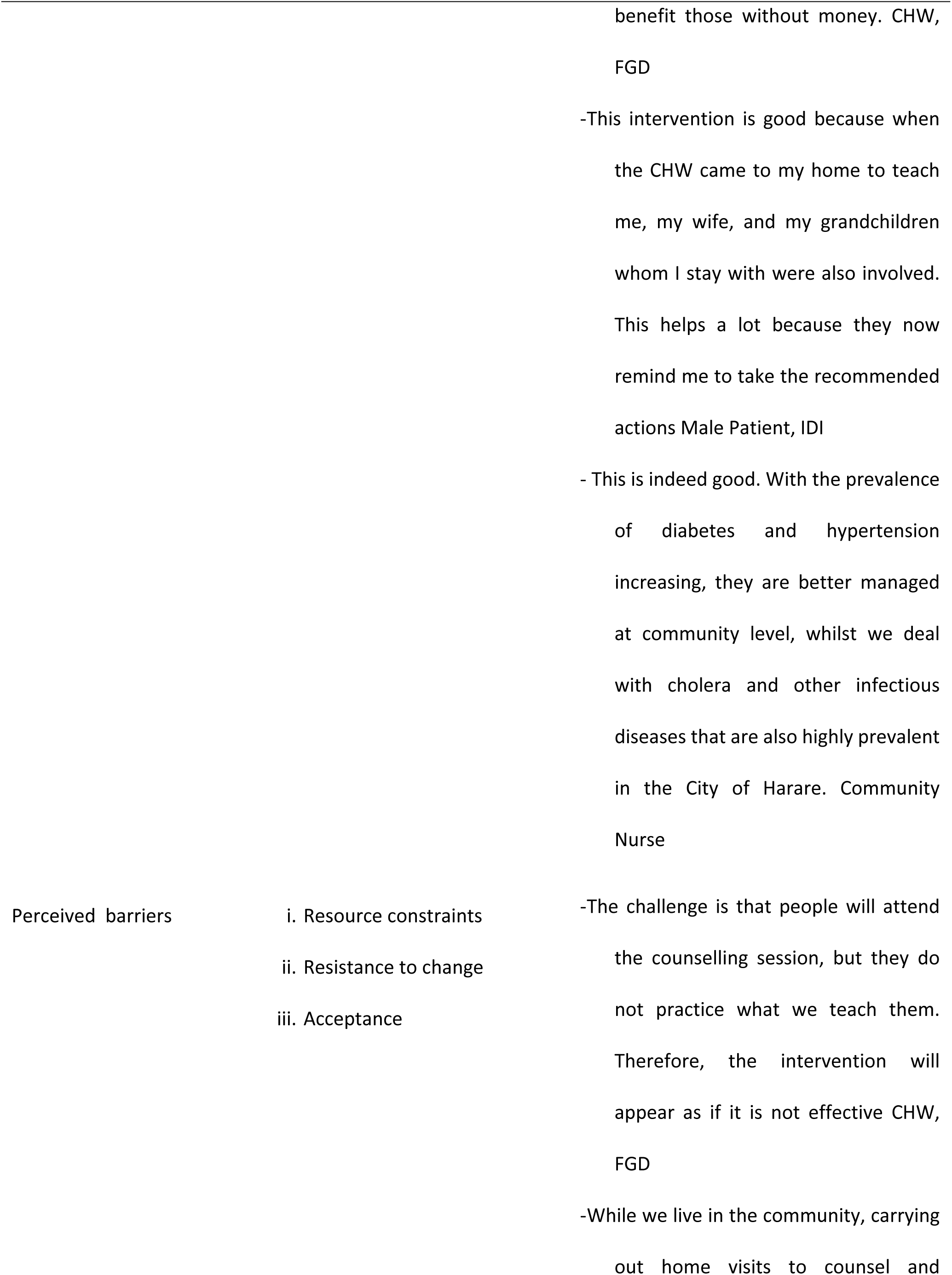
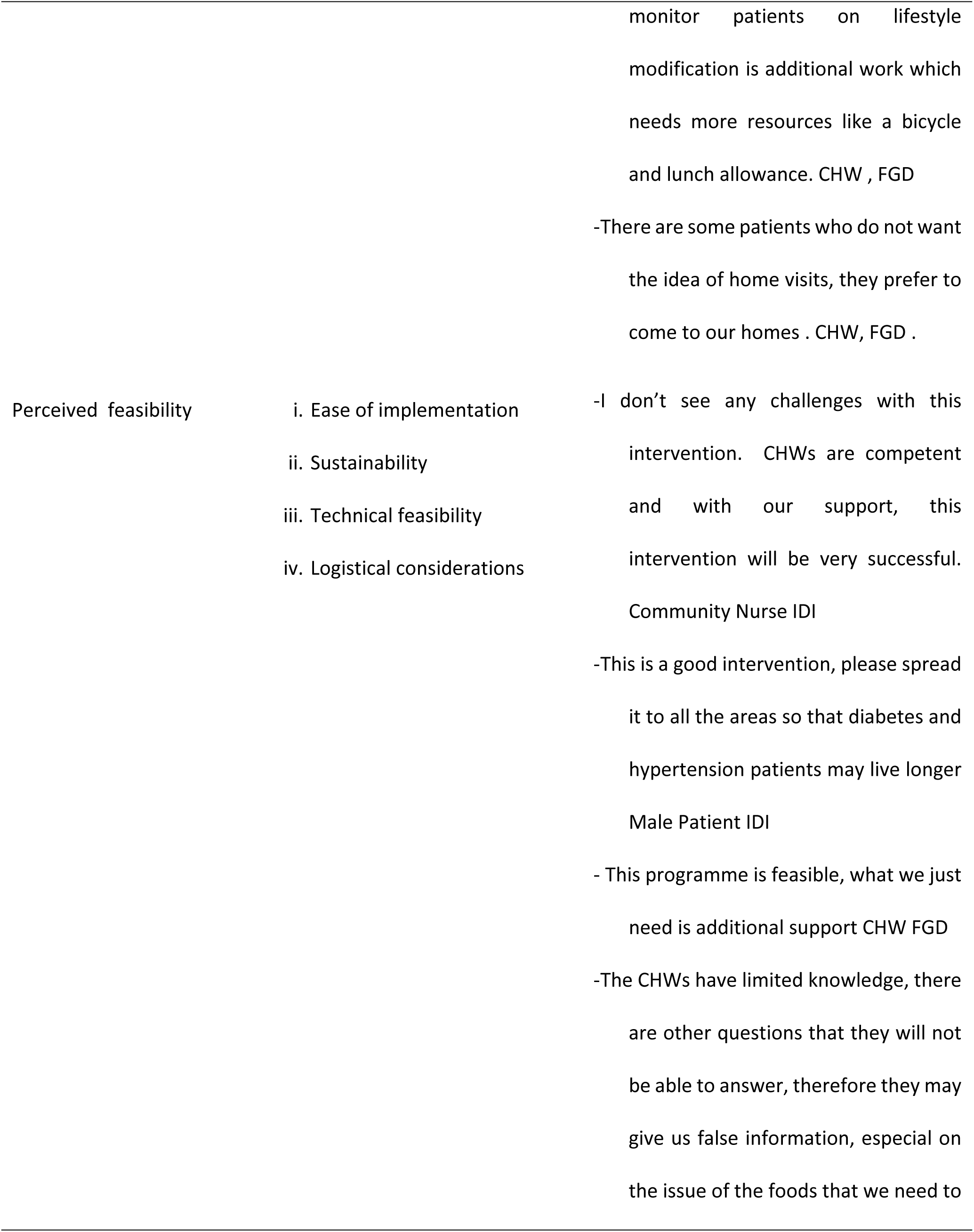
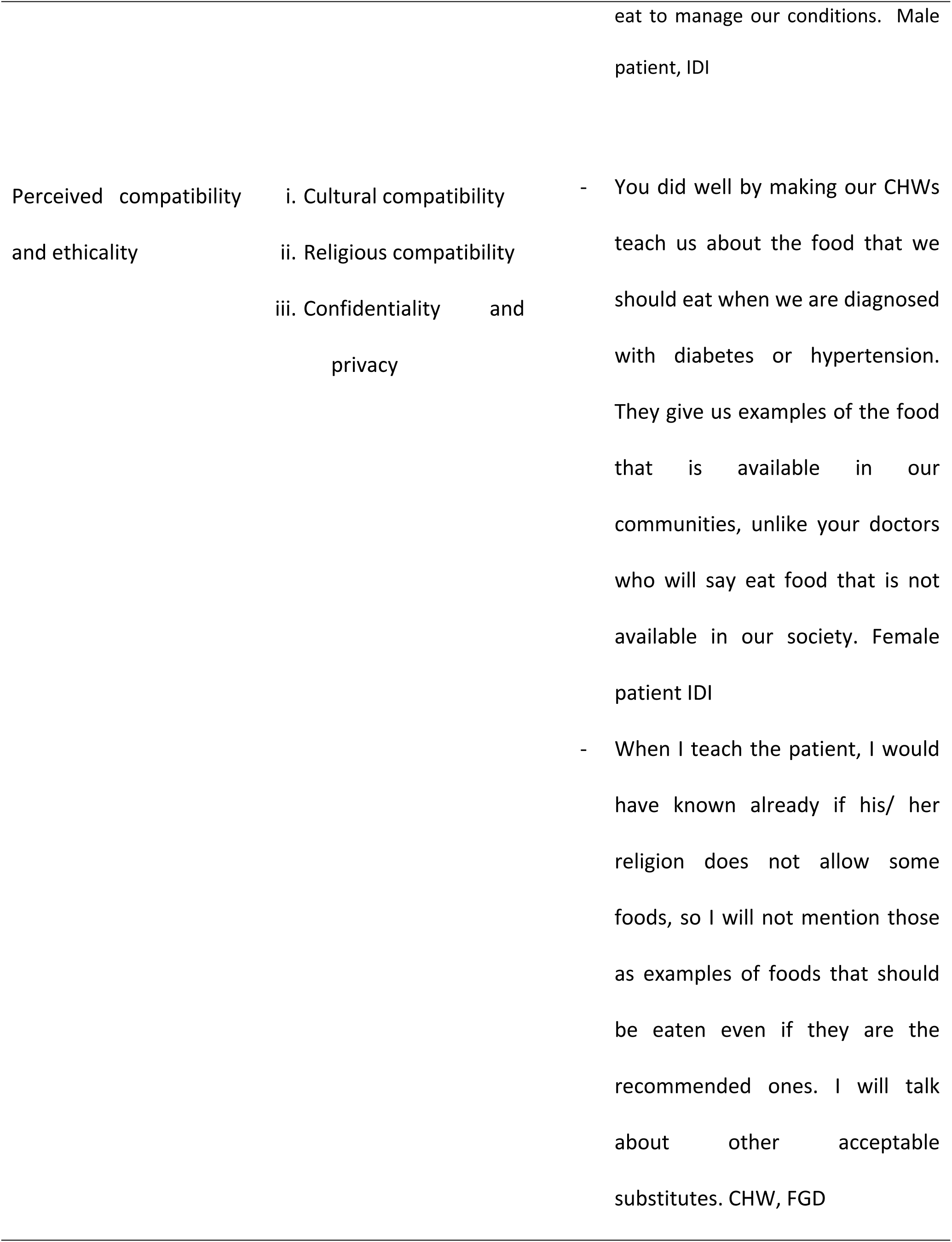
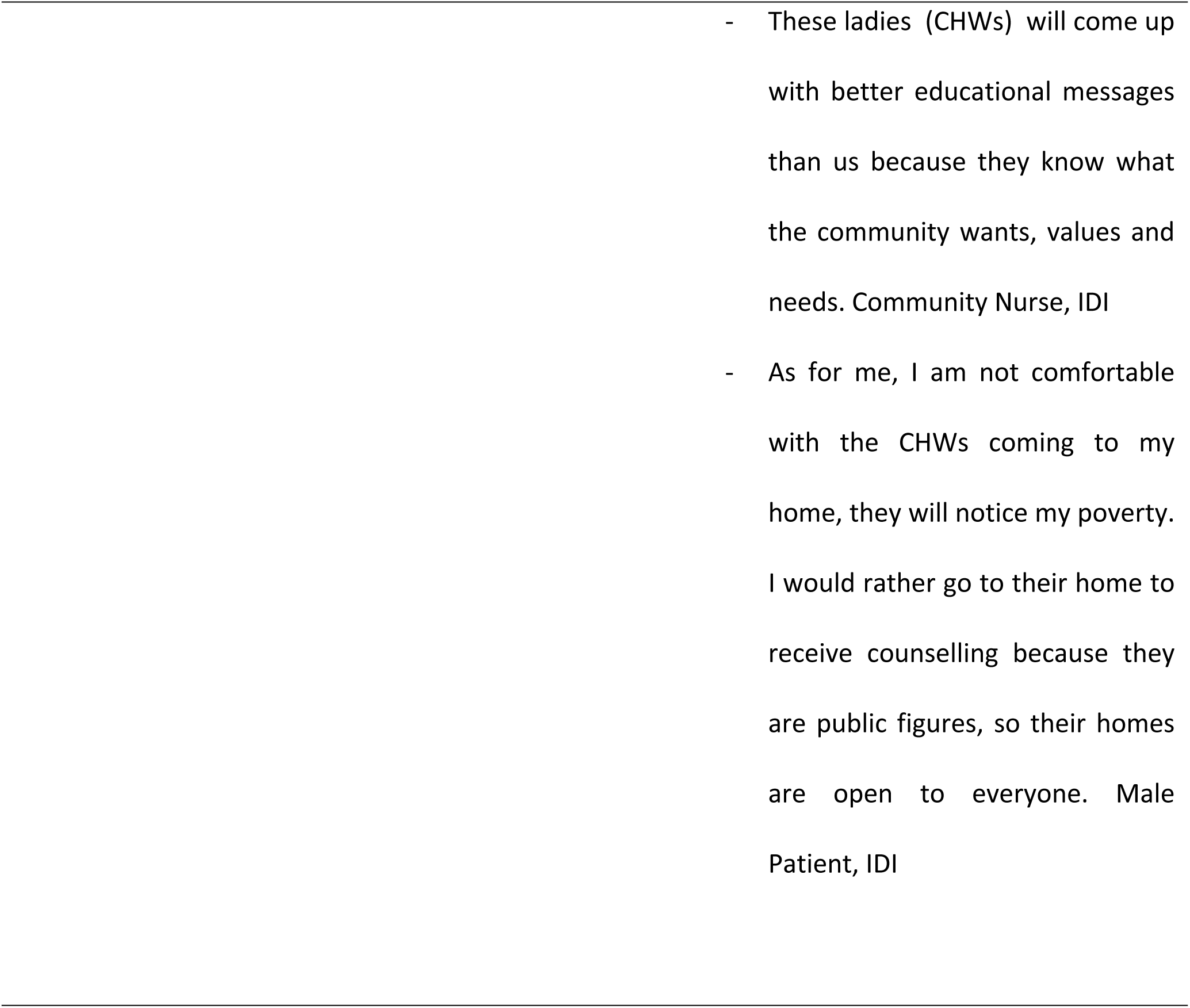
Emerging Themes, Sub-Themes and Supporting Verbatim

#### Perceived benefits

There was consensus that the intervention had many benefits amongst CHWs and community nurses. However, amongst patients, there were mixed perceptions regarding the benefits of the intervention.

Patients highlighted easy access to information on lifestyle modification and involvement of social support as the key benefits of the intervention. Community nurses also mentioned relief from high workload as another benefit of the intervention.

Easy access to information about lifestyle modifications required to manage hypertension and diabetes was the benefit that was mentioned mostly by the patients. All patients who were interviewed believed if CHWs are trained on the recommended lifestyle modifications, they will be able to get the information easily within their communities, without the need to visit the local clinics or private hospitals. Some of the patients also highlighted that they trust their community health workers and had a good relationship with them such that it was easier to get information without any fear. One of the patients (elderly female) commented that *“this intervention is very good. It will make us save money. We will no longer need to go to the clinics to ask about how to manage our conditions”.* Another male patient also commented that *what’s good about your intervention is that the CHWs live with us, so we can get information from them anytime, even on Sundays when you don’t open your clinics”*.

From the perspective of CHWs, the benefits that were mentioned include increased access to health care, improved quality of life of diabetes and hypertensive patients and cost effectiveness. The CHWs believed this intervention has the benefit of serving as a bridge between clinics and the patients in the community, thus improving access to health care for diabetes and hypertension patients who are mostly of advanced age. One CHW commented that “Our *patients are usually old and they do not have money to go to the clinics. So teaching them at their home is a big benefit”.* Another CHW also added that *“For diabetes and hypertensive patients, going to the clinic to get information only is a mammoth task because they need to pay consultation fee (USD5), Therefore this intervention will also benefit those without money”* Patients, particularly the male ones highlighted the involvement of other family members in the counselling sessions as a key benefit of the intervention. One of the male patients said “*This intervention is good because when the CHW came to my home to teach me, my wife, and my grandchildren whom I stay with were also involved. This helps a lot because they now remind me to take the recommended actions.”*

All nurses who were interviewed commended the intervention for reliving them from the duty of making home visits as well as counselling the patients. They believed that empowering the CHWs to counsel and monitor patients with regards to lifestyle modification is a good practice, considering that most health facilities in the City of Harare are short staffed. One of the nurses had this to say “*This is indeed good. With the prevalence of diabetes and hypertension increasing, they are better managed at community level, whilst we deal with cholera and other infectious diseases that are also highly prevalent in the City of Harare.”*

#### Perceived effectiveness

One of the benefits that was mostly mentioned across all participant groups was effectiveness of the intervention. All community health and the CHWs believed the intervention is effective in improving adherence to recommended lifestyle modifications and overall health outcomes among the patients. However, when it came to the patients, there were some who believed the intervention was not effective. One community nurse commented that ‘’ *Lifestyle modification is an important component of managing hypertension and diabetes, but some of our patients do not know that. Therefore, this intervention will help people to understand”*. A CHW also commented that “*teaching people is indeed helpful. We have done that in other programmes such as the Friendship bench. Those programmes were quite useful. Therefore, this intervention will also be effective*”. One of the patients *who did not believe that the intervention was not effective had this to say; “interventions that focus on teaching people do not work. Only medication helps sick people”*.

#### Perceived compatibility and ethicality

All categories of participants believed that since the intervention is implemented by CHWs, it will help in addressing cultural issues important to lifestyle modification, by virtue of sharing the same cultural and linguistic background with the patients. For instance, one of the patients said “*You did well by making our CHWs teach us about the food that we should eat when we are diagnosed with diabetes or hypertension. They give us examples of the food that is available in our communities, unlike your doctors who will say eat food that is not available in our society”.* Regarding the same issue, one of the CHWs also commented that *“when I teach the patient I would have known already if his/ her religion does not allow some foods, so I will not mention those as examples of foods that should be eaten even if they are the recommended ones. I will talk about other acceptable substitutes”.* A community nurse also said that “T*hese ladies (CHWs) will come up with better educational messages than us because they know what the community wants, values and needs”*. However, the CHWs also highlighted that there are some patients who are reluctant to be visited at their homes but prefer to meet the CHWs at other places such as the CHW’s home, the clinic or even at church because the want to maintain their privacy. This opinion was also raised by one of the patients who said *“as for me, I am not comfortable with the CHWs coming to my home, they will notice my poverty. I would rather go to their home to receive counselling because they are public figures, so their homes are open to everyone.”*

#### Perceived Barriers

Potential challenges of this intervention were mainly mentioned by the CHWs. The main challenges that were mentioned by CHWs include resistance to advice by patients, insufficient resources, and lack of acceptance at some of the patient’s homes. One of the CHWs said that “*the challenge is that people will attend the counselling session, but they do not practice what we teach them. Therefore, the intervention will appear as if it is not effective*”. Another CHW commented that “*whilst we live in the community, carrying out home visits to counsel and monitor patients on lifestyle modification is additional work which needs more resources like a bicycle and lunch allowance”*.

#### Perceived feasibility

Regarding feasibility of implementing the intervention, there were mixed perceptions, with community nurses and CHWs believing that it was feasible to implement the intervention whilst some of the patients were a bit sceptical. Patients believed whilst the CHWs were trained to implement this intervention, their training is limited and that can affect the quality of advice that they give to patients. The patients went on to highlight that the nurses need to monitor and support these CHW. One of the female patients said this regarding that “*the CHWs have limited knowledge, there are other questions that they will not be able to answer, therefore they may give us false information, especial on the issue of the foods that we need to eat to manage our conditions.”*

Community health nurses did not highlight any potential challenges with this intervention. One of the nurses said, “*I don’t see any challenges with this intervention. CHWs are competent and with our support, this intervention will be very successful”*. This was also supported by other patients. One of the patients said that “*this is a good intervention, please spread it to all the areas so that diabetes and hypertension patients may live longer*.” A CHW said *this programme is feasible, what we just need is additional support*”.

## DISCUSSION

As the burden of NCDs continue to rise worldwide, LMICs continue to struggle to manage these conditions due to several challenges, that include staff shortages[26]. One emerging global strategy to address staff shortages in management of chronic conditions is using CHWs to reach vulnerable populations in under-resourced settings[27]. Several reviews have found that over 60% of CHWs in LMIC performed some sort of home-based care; however, most of these efforts focused on HIV/AIDS, immunization and cancer and their work has also been mainly in rural areas[28,29]. Expanding on this work, we conducted a cluster randomized to assess the effects of community health worker -led health literacy intervention for lifestyle modification among hypertensive and diabetes patients that we designed[24] . To enhance our assessment of the intervention, we embedded a qualitative acceptability study on this cluster randomized trial. Our study aimed to explore the acceptability of a community health worker-led health literacy intervention on lifestyle modification among hypertension and diabetes patients in the City of Harare.

Findings from our study indicated that, the intervention is generally acceptable by the patients, the CHWs and community nurses, based on five dimensions, which are perceived benefits, perceived barriers, perceived compatibility, perceived feasibility and perceived effectiveness. Interventions involving community health workers have been proven to be acceptable in other settings world-wide [22,30]. This could be because CHWs are respectable community members, hence their advice can be easily accepted within communities. The fact that they undergo some training also enhances their acceptability in the communities that they serve. Training was highlighted as the key facilitators of acceptability of CHWs in a rural area in South Africa [2].

Whilst our intervention was perceived to have many benefits by all the categories of participants (patients, CHWs and nurses), it is also important to take note of the potential challenges that were mentioned by the patients and the CHWs. Patients were concerned that the CHWs may not be competent to respond to some of their questions. This is an important finding which should not be ignored when scaling up the intervention. One strategy of addressing this concern is through thorough training of CHWs before implementing the intervention and close support and supervision by nurses during implementation. As highlighted by Stansert et al, CHWs are accepted when the community know that they are trained and when they receive occasional support from professional healthcare workers [2] The CHWs themselves also highlighted the need for extra support in terms of transport and financial allowances. This should also be considered when scaling up the intervention. Inadequate resources has been highlighted as one of the key challenges that hinder effective implementation of CHW interventions in LMICs[5,30,31], hence in needs to be addressed to ensure programme success.

Our study has several notable strengths. We conducted the study during the implementation of the intervention. Therefore, recall bias was minimized since the participants were still recalling their experience with the implementation of the intervention. We also assessed the acceptability from the perspectives of all those categories of people who are key to the success of the intervention (the beneficiaries-patients, the implementers-CHWs and the supervisors-community nurses). Therefore, our findings are quite rich. Nevertheless, there are some limitations to our study. We cannot rule out the possibility of social desirability bias, particularly from the patients and the CHWs. There is a possibility that they could not have highlighted the potential challenges of the intervention because the principal investigator was involved in the conduct of the in-depth interviews and the focus group discussions. We tried to minimize this by limiting the role of the principal investigator to recording responses only. Two research assistants who were not part of the intervention are the ones who interviewed participants and facilitated focus group discussions. Secondly, results on the perceived effectiveness could be limited because the data was collected during the implementation of the intervention, such that the ultimate benefit may not have been experience. To this end, we recommend a post acceptability assessment. Nonetheless, the results for this study provide valuable information for upscaling of the intervention.

## CONCLUSION

Although there is vast evidence regarding effectiveness of community-based interventions for NCDs, effectiveness alone does not guarantee the success of public health interventions. Acceptability is also important, because many effective interventions may not yield the expected results due to low uptake resulting from unacceptable interventions. Our community health worker-led health literacy intervention for lifestyle modification intervention was perceived as having many benefits and thus acceptable by hypertension and diabetes patients, community health workers and community nurses in the city of Harare.

## ACKNOWLEDGEMENTS

We would like to acknowledge the City of Harare, City Health Director for granting permission to carry out the study in the city of Harare health facilities. We also extent our gratitude to the study participants (diabetes and hypertension patients, CHWs and community nurses). We would also like to thank our colleagues from the department of global public health and family medicine for per review of our study proposal Research reported in this publication was financially supported by the Fogarty International Center and National Institute of Dental and Craniofacial Research of the National Institutes of Health under Award Number D43 TW011968. The content is solely the responsibility of the authors and does not necessarily represent the official views of the National Institutes of Health

## DATA AVAILABILITY

The consent and ethical approvals for this study does not have any restrictions for data sharing. Therefore, the data sets generated from this study will be made available to the public upon request.

## AUTHORS’ CONTRIBUTIONS

NAK: conceptualisation of study, data collection, data analysis and drafting of first draft and subsequent drafts leading to the final manuscript; ED and AD; data collection, data analysis and reviewing of initial manuscript and subsequent drafts . SR, SS and GTF: Reviewing of study proposal, reviewing of initial manuscript and subsequent drafts. All authors read and approved the final manuscript.

